# Interleukin-6 as a predictor of early weaning from invasive mechanical ventilation in patients with acute respiratory distress syndrome

**DOI:** 10.1101/2022.04.04.22273418

**Authors:** Kazuya Sakai, Mototsugu Nishii, Ryo Saji, Reo Matsumura, Fumihiro Ogawa, Ichiro Takeuchi

## Abstract

**Background:** Therapeutic effects of steroids on acute respiratory distress syndrome (ARDS) requiring mechanical ventilation (MV) have been reported. However, predictive indicators of early weaning from MV post-treatment have not yet been defined, making treating established ARDS challenging. Interleukin (IL)-6 has been associated with the pathogenesis of ARDS.

**Objective:** Our aim was to clarify clinical utility of IL-6 level in ventilated patients with established ARDS.

**Methods:** Clinical, treatment, and outcome data were evaluated in 119 invasively ventilated patients with severe acute respiratory syndrome coronavirus 2 (SARS-CoV-2)-mediated ARDS. Plasma levels of IL-6 and C-reactive protein (CRP) were measured on days 1, 4, and 7 after intubation.

**Results:** Fifty-two patients were treated with dexamethasone (steroid group), while the remaining 67 patients were not (non-steroid group). Duration of MV use was significantly shorter in the steroid group compared to non-steroid group (11.5±0.6 vs. 16.1±1.0 days, P = 0.0005, respectively) along with significantly decreased levels of IL-6 and CRP. Even when restricted to the steroid group, among variables post-MV, IL-6 level on day 7 was most closely correlated with duration of MV use (Spearman’s rank correlation coefficient [ρ] = 0.73, P < 0.0001), followed by CRP level on day 7 and the percentage change in IL-6 or CRP levels between day 1 and day 7. Moreover, among these variables, IL-6 levels on day 7 showed the highest accuracy for withdrawal from MV within 11 days (AUC: 0.88), with optimal cutoff value of 20.6 pg/mL. Consistently, the rate of MV weaning increased significantly earlier in patients with low IL-6 (≤ 20.6 pg/mL) than in those with high IL-6 (> 20.6 pg/mL) (log-rank test P < 0.0001).

**Conclusions:** In invasively ventilated patients with established ARDS due to SARS-CoV-2, plasma IL-6 levels served as a predictor of early withdrawal from MV after dexamethasone administration.

## Introduction

Acute respiratory distress syndrome (ARDS), characterized by acute diffuse inflammatory lung injury and rapidly impaired oxygenation requiring mechanical ventilation (MV), represents a common pulmonary disorder in intensive care unit (ICU). ARDS can be caused by many etiologies, such as trauma, transfusion history, infection, sepsis, pneumonia, and even ventilator-induced lung injury.^1-3^ Moreover,

in-hospital mortality remains high at approximately 40-50% in overall patients with ARDS, and further increases along with severity of ARDS, from mild to severe.^4-5^ Against this background, development of underlying therapies for ARDS is needed. So far, a number of randomized controlled studies have been conducted to evaluate the efficacy of corticosteroids with the ability to modulate hyperinflammation on ARDS.^6-10^ Based on these observations, the effectiveness of corticosteroids on ARDS is still controversial ^8-9^ yet early administration of dexamethasone is likely to reduce duration of MV use and mortality in patients with moderate to severe ARDS.^10^ Recently, at the end of 2019, COVID-19 outbreak unexpectedly emerged from Wuhan, China, and quickly spread around the world.^11^ Critically ill COVID-19 patients who required MV suffered from ARDS caused by severe acute respiratory syndrome coronavirus 2 (SARS-CoV-2) and had a high in-hospital mortality.^12^ Consistent with previous report,^10^ early administration of dexamethasone as well as remdesivir, an antiviral agent was reported to reduce the duration of MV use and in-hospital mortality in patients with ARDS caused by SARS-CoV-2.^13-14^ However, the treating ARDS with steroid therapy remains challenging, because the fundamental indicators to predict early withdrawal from MV post-treatment have not yet been defined. Studies with experimental models of ARDS have demonstrated that that lung injury can be induced by primed alveolar macrophages.^15-16^ Consistently, the autopsy and pathological studies of SARS-CoV-2-induced ARDS patients suggested that over-activation of alveolar macrophages causes cytokine storm, resulting in severe damage to lung tissue.^17^ Interleukin (IL)-6, derived from inflammatory monocytes and alveolar macrophage, may be responsible for severe lung inflammation and pulmonary function disability in the pathophysiology of ARDS. It has been reported that in the bronchoalveolar lavage fluid (BALF) from patients at risk for ARDS or with established ARDS, which mainly contains inflammatory monocytes and alveolar macrophages, IL-6 levels were extremely increased and remained high throughout the course of ARDS.^18-20^ Moreover, detailed single-cell RNA sequencing data obtained from BALF and peripheral blood mononuclear cells from patients with SARS-CoV-2-induced ARDS have recently shown that IL-6 is released into the systemic circulation from SARS-CoV-2-affected lungs, rather than from peripheral immune cells.^21-22^ Thus, we hypothesized that circulating IL-6, which reflects disease activity, may be a predictive indicator of early withdrawal from MV in patients with established ARDS.

To facilitate treatment of established ARDS, we evaluated this hypothesis in critically ill patients with SARS-CoV-2-mediated ARDS.

## Materials and methods

### Study design and setting

This was a single retrospective observational study in Japan. COVID-19 patients admitted to the Yokohama City University Hospital (YCUH) between February 2020 and September 2021 were enrolled in this study. Enrolled patients were observed until hospital discharge after the enrollment to evaluate clinical outcome. Discharge was determined after the patient was weaned from invasive MV. This study followed the Transparent Reporting of a multivariable prediction model for Individual Prognosis or Diagnosis reporting statement for prognostic studies.

### Study sample

The inclusion criteria were as follows: (1) diagnosis of SARS-CoV-2-induced ARDS by reverse transcription-polymerase chain reaction (RT-PCR) assay for SARS-CoV-2, chest computed tomography (CT) scan, and the ratio of the partial pressure of arterial oxygen to fraction of inspired oxygen (P/F ratio); (2) more than 18 years; (3) the use of invasive MV corresponded to the need for intubation and intensive care unit (ICU) management. Moreover, ARDS was diagnosed according to the Berlin definition.^23^ Invasive MV was introduced if P/F ratio was less than 100 under oxygen support from a reservoir mask at 8 L/min. The exclusion criteria were as follows (Fig. S1): (1) patients not requiring invasive MV; (2) those with missing data, including clinical, laboratory, and outcome data; (3) those without consent for participation.

Ultimately, SARS-CoV-2-induced severe ARDS patients requiring invasive MV with complete clinical, laboratory, and outcome data as well as consent for participation were evaluated in the final analysis.

### Management of ventilated patients with ARDS

Management of ARDS and weaning from MV were performed according to the guideline for the management of ARDS in Japan.^24^

### Ethical considerations

This study was approved by the Institutional Ethics Board of the YCUH (No. B210100010). During hospitalization, patients were provided negative and positive information regarding this study, including the purpose and contribution of this study, the use of personal information, and complications associated with blood collection, and were asked to participate in this study. Ultimately, we obtained written informed consent for participation in the study and access to medical and laboratory records from patients. Alternatively, we were unable to obtain written consent for participation from patients who died without being weaned after being placed on a ventilator, which means that we used an opt-out method to obtain consent for this study from them. The study had no risk/negative consequence on those who participated in the study. Medical record numbers were used for data collection and no personal identifiers were collected or used in the research report. Data was accessed from February 16, 2020, to October 30, 2021, and access to the collected information was limited to the principal investigator and confidentiality was maintained throughout the project.

### Data and Specimen collection

Clinical data and treatment and outcome data were obtained from electronic medical records. Two researchers independently reviewed the data collection forms to double-check the collected data.

### IL-6 and C-reactive protein (CRP) measurements

Plasma levels of IL-6 and CRP were measured on days 1, 4, and 7 after intubation. The fully automated Elecsys system on a cobas e801 platform (Roche Diagnostic GmBH, Mannheim, Germany) was used for IL-6 measurement. CRP levels were measured on a cobas c702 platform by using the Tina-quant CRP assay (Roche Diagnostic GmBH, Mannheim, Germany).

### Statistical analysis

Data analyses were done using the JMP ver. 12.2 software. All categorical variables were presented as number (n), %. Continuous variables are shown as mean±SEM and 95% confidence interval [CI]. Differences between the different treatment groups were analyzed with Fisher’s exact test (for categorical data) or the Mann-Whitney *U* test (for continuous data). Logistic regression analysis was used for univariate analysis of IL-6 and CRP levels. Correlations between variables were evaluated by the Spearman’s rank test. To evaluate the area under curves (AUCs) and cutoff values of parameters, the receiver operator characteristic (ROC) curves were constructed. The optimal cutoff was defined as value which had the best compromise between sensitivity and specificity for predicting withdrawal from MV on ROC curve. Comparisons of ventilator weaning rates between subgroups were performed with the Log-rank test. Statistical significance was set at P < 0.05.

## Results

### Baseline characteristics

From February 2020 to July 2021, 268 patients with COVID-19 pneumonia were enrolled. Of those, 149 patients not using invasive MV or without complete data or consent for participation were excluded. Ultimately, a total of 119 invasively ventilated patients with SARS-CoV-2-induced ARDS were evaluated in this study (Fig. S1). Table 1 shows individual baseline clinical and outcome data in the present study population. The age was 63.1±1.2 years. Of these patients, 96 (81%) were male and 23 (19%) were female. The mean duration from symptom onset to intubation was 7.0 days. Approximately 20 to 30% had comorbidity such as chronic kidney disease, diabetes mellitus, and hypertension. Among clinical and laboratory data such as systolic blood pressure (SBP), platelet count, total bilirubin, and creatinine at inclusion, creatinine levels were elevated in some cases. Thus, no high-risk patients with sequential organ failure assessment scores of 5 or higher were identified in the study population. All patients received anticoagulation therapy. Almost patients (83%) were treated with remdesivir. In 44% (n = 52), treatment with dexamethasone (6 mg/day) was initiated before (n = 12) or on (n = 40) admission to YCUH and continued at least during MV use. Duration of MV use in overall patients was 13.9±0.6 days. Ultimately, 13 patients required ECMO or died of ARDS during hospitalization.

**Table1.**
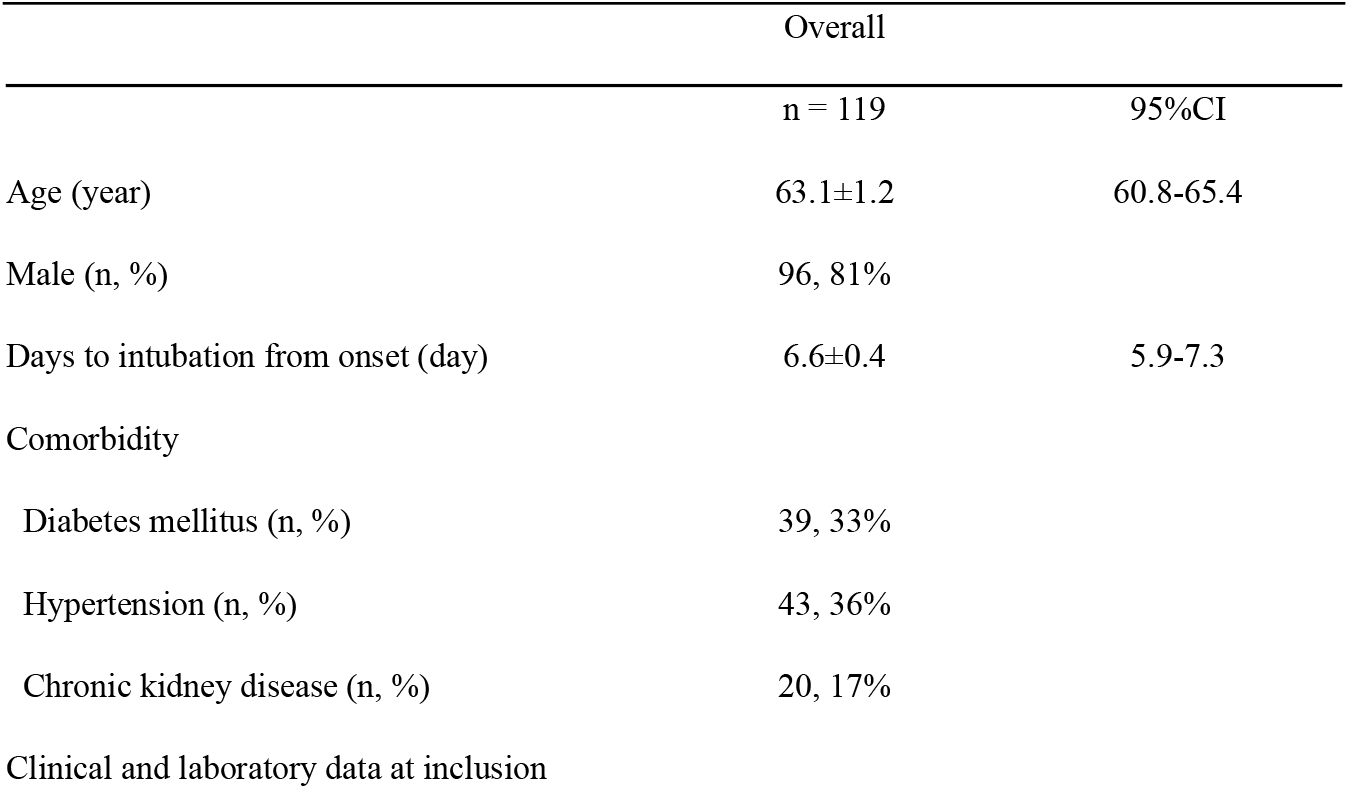
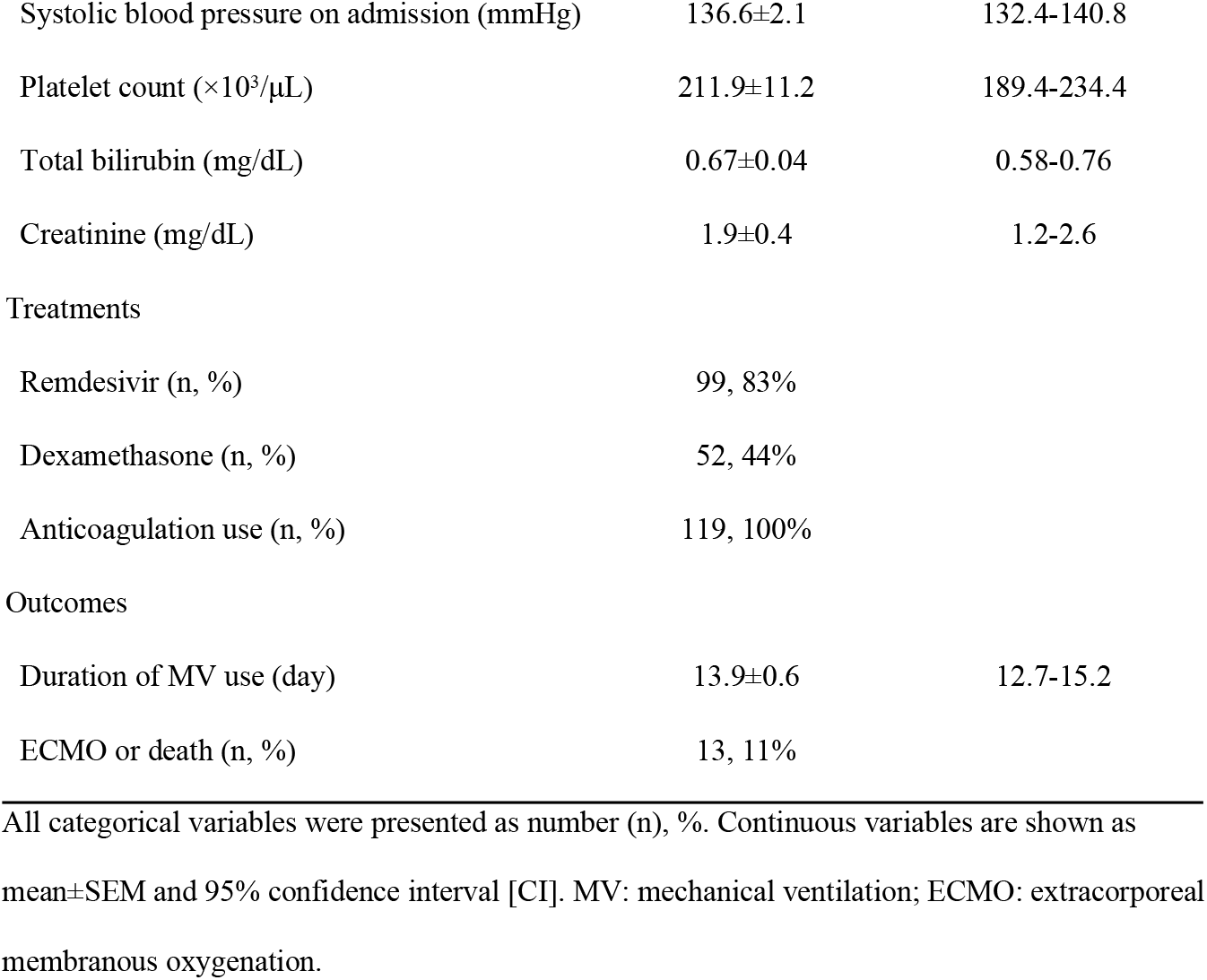
Clinical Characteristics.

|  | Overall |  |
| --- | --- | --- |
|  | n = 119 | 95%CI |
| Age (year) | 63.1±1.2 | 60.8-65.4 |
| Male (n, %) | 96, 81% |  |
| Days to intubation from onset (day) | 6.6±0.4 | 5.9-7.3 |
| Comorbidity |  |  |
| Diabetes mellitus (n, %) | 39, 33% |  |
| Hypertension (n, %) | 43, 36% |  |
| Chronic kidney disease (n, %) | 20, 17% |  |
| Clinical and laboratory data at inclusion |  |  |
| Systolic blood pressure on admission (mmHg) | 136.6±2.1 | 132.4-140.8 |
| Platelet count ($\times 10^3/\mu\text{L}$) | 211.9±11.2 | 189.4-234.4 |
| Total bilirubin (mg/dL) | 0.67±0.04 | 0.58-0.76 |
| Creatinine (mg/dL) | 1.9±0.4 | 1.2-2.6 |
| Treatments |  |  |
| Remdesivir (n, %) | 99, 83% |  |
| Dexamethasone (n, %) | 52, 44% |  |
| Anticoagulation use (n, %) | 119, 100% |  |
| Outcomes |  |  |
| Duration of MV use (day) | 13.9±0.6 | 12.7-15.2 |
| ECMO or death (n, %) | 13, 11% |  |

All categorical variables were presented as number (n), %. Continuous variables are shown as mean±SEM and 95% confidence interval [CI]. MV: mechanical ventilation; ECMO: extracorporeal membranous oxygenation.

### The effects of dexamethasone on clinical outcomes of invasively ventilated ARDS patients

To evaluate the efficacy of dexamethasone in invasively ventilated patients with SARS-CoV-2-induced ARDS, clinical outcomes as well as clinical characteristics were compared between patients who received dexamethasone therapy and those who did not (steroid group [n = 52] and non-steroid group [n = 67], respectively) (Fig. S1, Table 2). There was fewer elderly male in the steroid group than in the non-steroid group. Duration from symptom onset to intubation was significantly shorter in the steroid group than in the non-steroid group. Regarding with comorbidity, diabetes mellitus was fewer in the steroid group. Consideration of complications to steroids, including secondary infections and hyperglycemia, may have influenced differences of age and comorbidity between the 2 groups. Among clinical and laboratory data at inclusion, SBP was significantly lower in the steroid group than in the non-steroid group. However, no patients with SBP below 70 mmHg were identified in both groups. Platelet count, total bilirubin level, and creatinine level were not significantly different between the 2 groups. Anti-viral agent use had no significant difference between the 2 groups. Ultimately, combined event rate of ECMO use and in-hospital death was significantly lower in the steroid group than in the non-steroid group. Moreover, duration of invasive MV use was significantly shorter in the steroid group than in the non-steroid group. These results suggest that dexamethasone positively affects clinical outcomes in invasively ventilated patients with established ARDS caused by SARS-CoV-2.

**Table 2.**
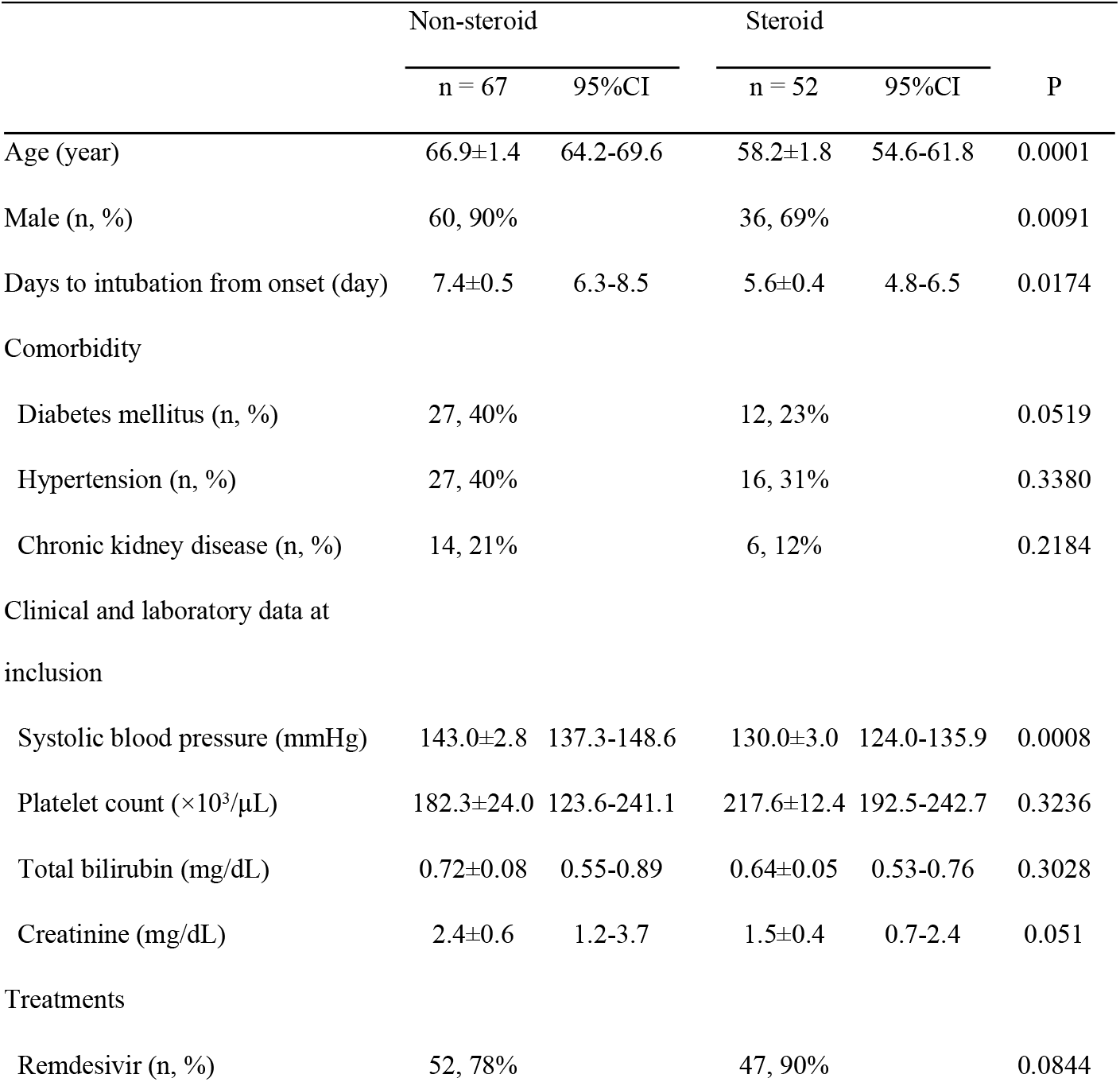
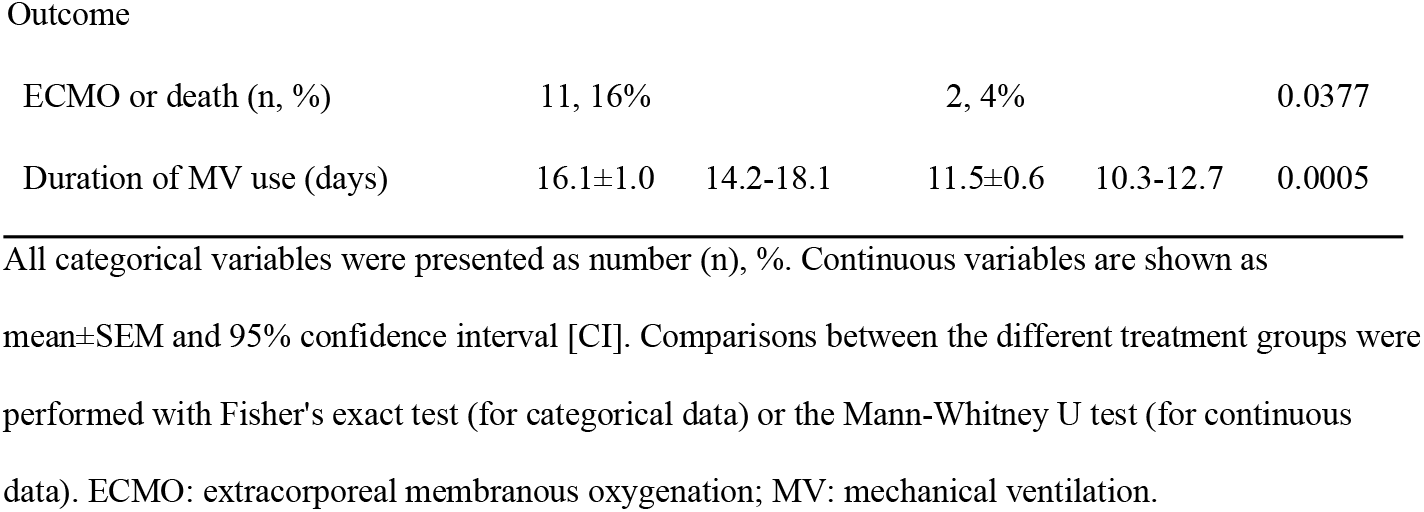
Comparisons of clinical and outcome data.

### The effect of dexamethasone in plasma levels of IL-6 and CRP

We evaluated if and how dexamethasone affects levels of IL-6 and CRP. In 56 patients (steroid group: n = 40 and non-steroid group: n = 16), plasma levels of IL-6 and CRP were measured on days 1, 4, and 7 after MV introduction. As shown in Table 3, levels of IL-6 and CRP were significantly lower in the steroid group compared to the non-steroid group. Collectively, dexamethasone decreased circulating levels of IL-6 as well as CRP in invasively ventilated patients with SARS-CoV-2-mediated ARDS.

**Table 3.** Comparisons of plasma levels of interleukin (IL)-6 and C-reactive protein (CRP) on days 1, 4, and 7 after mechanical ventilation introduction.

|  | Non-steroid |  | Steroid |  |  |
| --- | --- | --- | --- | --- | --- |
|  | n = 16 | 95%CI | n = 40 | 95%CI | P |
| IL-6 (pg/mL) |  |  |  |  |  |
| day 1 | 424.8±116.7 | 164.8-684.9 | 77.1±21.2 | 34.2-120.1 | < 0.0001 |
| day 4 | 413.7±60.8 | 281.3-546.1 | 57.6±21.4 | 14.0-101.2 | < 0.0001 |
| day 7 | 260.8±140.2 | -44.7-566.3 | 48.0±12.6 | 22.3-73.7 | 0.0002 |
| CRP (mg/dL) |  |  |  |  |  |
| day 1 | 19.6±3.1 | 12.9-26.3 | 8.6±0.9 | 6.7-10.5 | 0.0004 |
| day 4 | 23.2±2.0 | 18.8-27.6 | 6.7±1.3 | 3.9-9.4 | < 0.0001 |
| day 7 | 18.4±2.5 | 13.0-23.7 | 5.8±0.9 | 4.0-7.6 | < 0.0001 |

Continuous variables are shown as mean±SEM and 95% confidence interval [CI]. Comparisons between the different treatment groups were performed with the Mann-Whitney U test.

### Relationship between IL-6 levels and invasive MV duration in the presence of steroid therapy

We evaluated if IL-6 and CRP levels on days 1, 4, and 7 after intubation, the 2-day difference, or percentage of change were associated with duration of MV use in the steroid group. Among these variables, IL-6 level on day 7 was most closely correlated with MV duration (Spearman’s rank correlation coefficient [ρ] = 0.7253, P < 0.0001), followed by the percentage of change in CRP or IL-6 levels between day 1 and day 7 (ρ =0.4529, P = 0.0063; ρ = 0.3754, P = 0.0220; respectively), CRP level on day 7 (ρ = 0.3752, P = 0.0203), and IL-6 level on day 1 (ρ = 0.3668, P = 0.0255), while IL-6 level on day 4, CRP levels on day 1 and day 4, and difference of IL-6 or CRP levels between day 1 and day 7 were not significantly correlated (Table 4 and Fig. 1). These results suggested that IL-6 as well as CRP after intubation is an indicator of duration of MV use in SARS-CoV-2-mediated ARDS patients who were treated with dexamethasone.

**Table 4.** The relationship between plasma levels of interleukin (IL)-6 and duration of invasive mechanical ventilation (MV) use.

| Variables | $\rho$ | P |
| --- | --- | --- |
| IL-6 day 1 | 0.3668 | 0.0255 |
| IL-6 day 4 | 0.3067 | 0.0731 |
| IL-6 day 7 | 0.7253 | < 0.0001 |
| 2-day difference in IL-6 | 0.1866 | 0.2760 |
| % change in IL-6 | 0.3754 | 0.0220 |
| CRP day 1 | 0.0286 | 0.8611 |
| CRP day 4 | 0.2848 | 0.1025 |
| CRP day 7 | 0.3752 | 0.0203 |
| 2-day difference in CRP | 0.3155 | 0.0648 |
| % change in CRP | 0.4529 | 0.0063 |

Correlations between plasma levels of IL-6 and C-reactive protein (CRP) on days 1, 4, and 7 after the introduction of MV and duration of MV use in the steroid group (n = 40) were analyzed with Spearman's rank correlation coefficient ( $\rho$ ). 2-day difference and percentage (%) of change in IL-6 or CRP were calculated as follows: level on day 7 - level on day 1; 2-day difference / level on day 1 × 100%; respectively.

**Fig 1.**
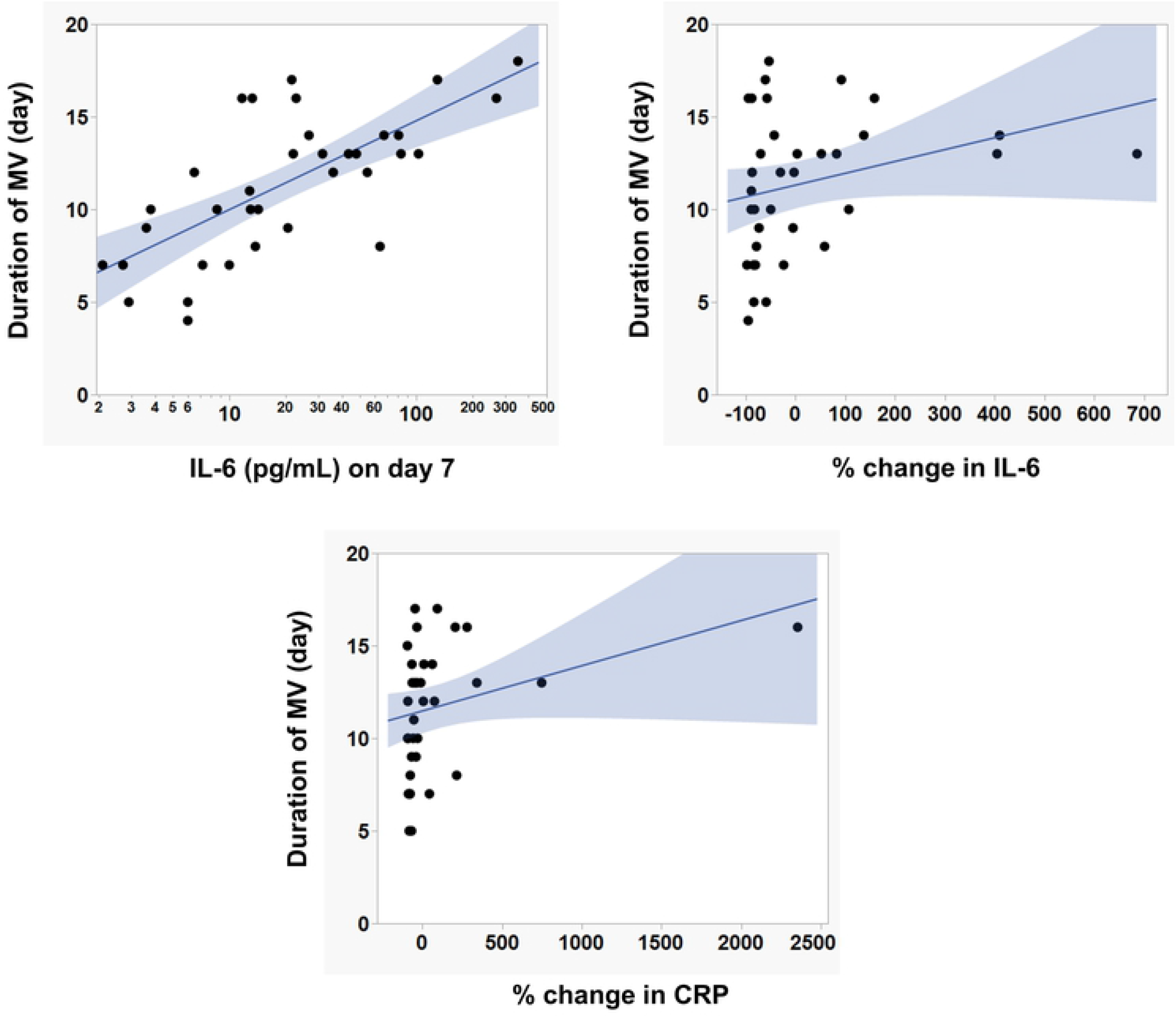
The relationship between interleukin (IL)-6 levels and duration of invasive mechanical ventilation (MV) use. In invasively ventilated patients with SARS-CoV-2-mediated ARDS who were treated with dexamethasone (n = 40), plasma levels of IL-6 on day 7 after intubation and percentage of change in IL-6 or CRP between day 1 and day 7 were correlated with duration of MV use with Spearman’s rank test. Closed dots indicate individual data.

### Association between IL-6 level and early withdrawal of invasive MV after steroid administration

The ROC analysis in the steroid group showed that among significantly correlated variables with MV duration, IL-6 level on day 7 after intubation was most strongly predictive of MV withdrawal within 11 days (median value of MV duration in the steroid group) (AUC: 0.88 [95%CI 0.72-0.96], P = 0.0001), followed by the percentage of change in CRP or IL-6 level between day 1 and day 7 (AUC: 0.78 [95%CI 0.58-0.90], P = 0.0149; AUC: 0.73 [95%CI 0.54-0.86], P = 0.0051; respectively) and CRP level on day 7 (AUC: 0.76 [95%CI 0.56-0.89], P = 0.0024) (Table 5 and Fig. 2). The optimal cut-off value of IL-6 on day 7 was 20.6 pg/mL, with sensitivity of 88% and specificity of 84% (Table 5 and Fig. 2). IL-6 was predictive of earlier withdrawal from invasive MV after steroid administration in intubated patients with SARS-CoV-2-mediated ARDS.

**Table 5.**
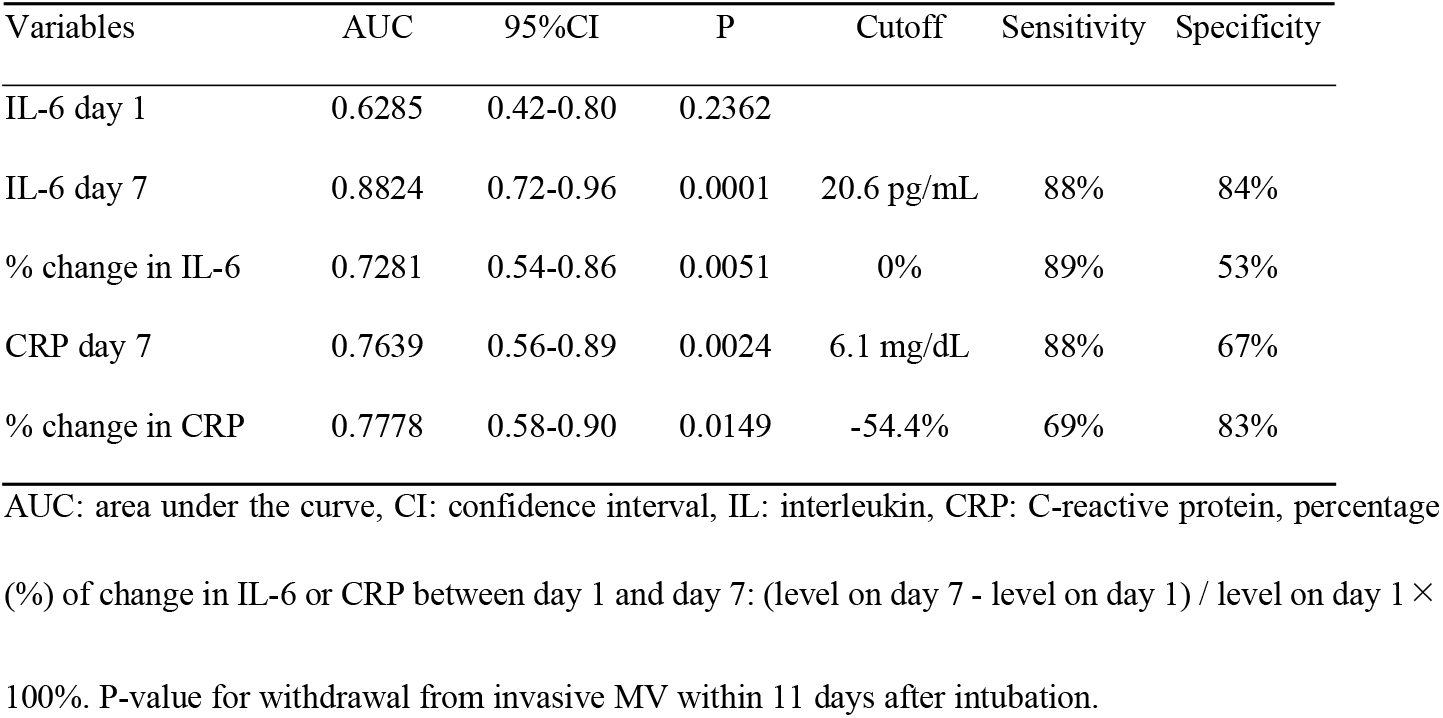
Receiver operating characteristic analyses for associations of different variables and withdrawal from invasive mechanical ventilation (MV)

**Fig 2.**
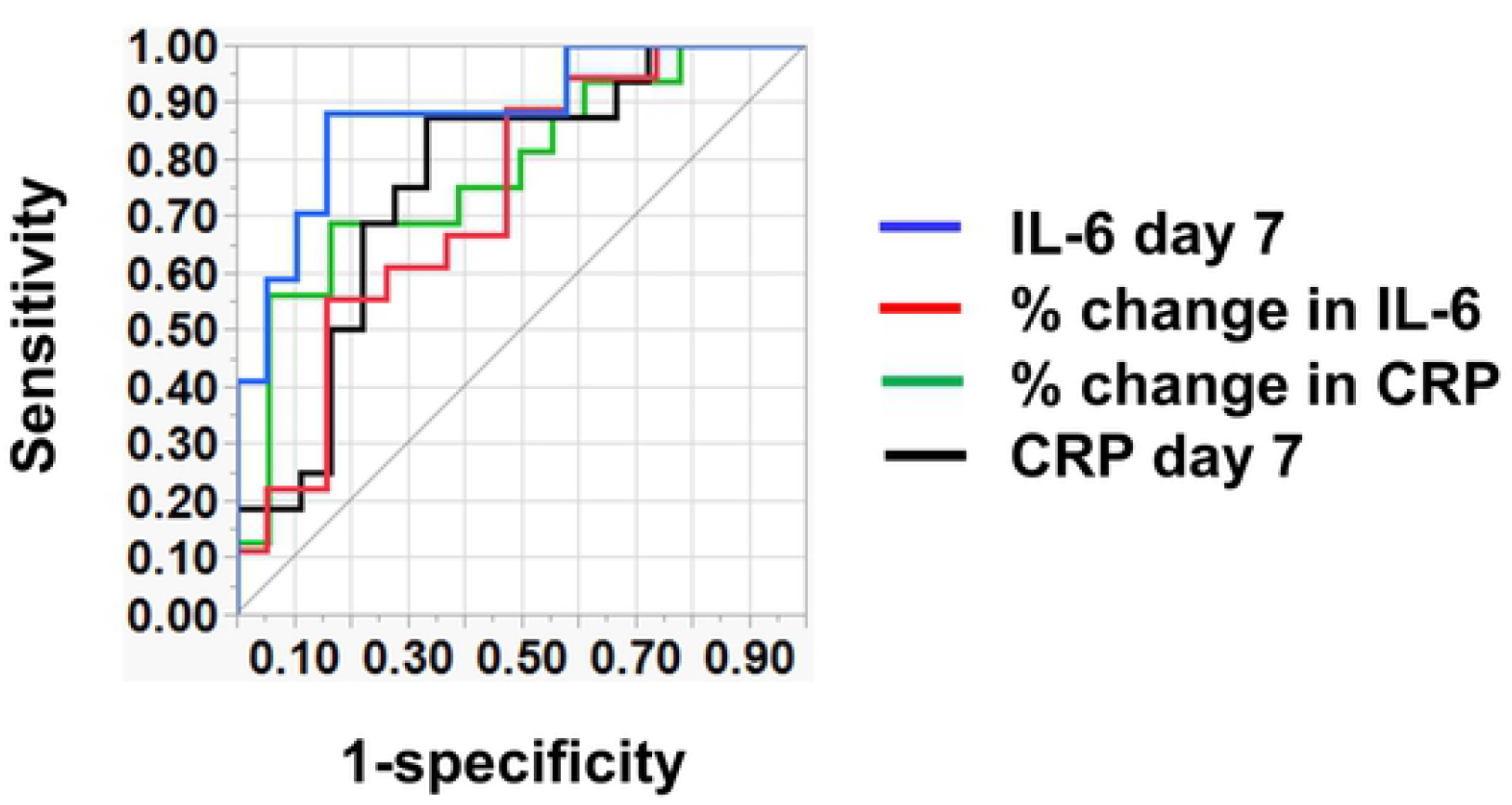
Receiver operating characteristic analysis for withdrawal from invasive mechanical ventilation (MV) The area under the receiver operating characteristic curve of the logistic regression models (interleukin [IL]-6 level on day 7 after intubation: blue; percentage [%] of change in IL-6 or CRP between day 1 and day 7: red or green, respectively; C-reactive protein [CRP] level on day 7 after MV introduction: black). % change in IL-6 or CRP was calculated as follow: (level on day 7 - level on day 1) / level on day 1× 100%).

Moreover, Kaplan-Meier curves in the steroid group that were constructed according to above or below optimal cutoffs demonstrated that the cumulative weaning rate from invasive MV increased significantly earlier in patients with low IL-6 (≤ 20.6 pg/mL, n = 19) compared to those with high IL-6 (> 20.6 pg/mL, n = 21) (Fig. 3).

**Fig 3.**
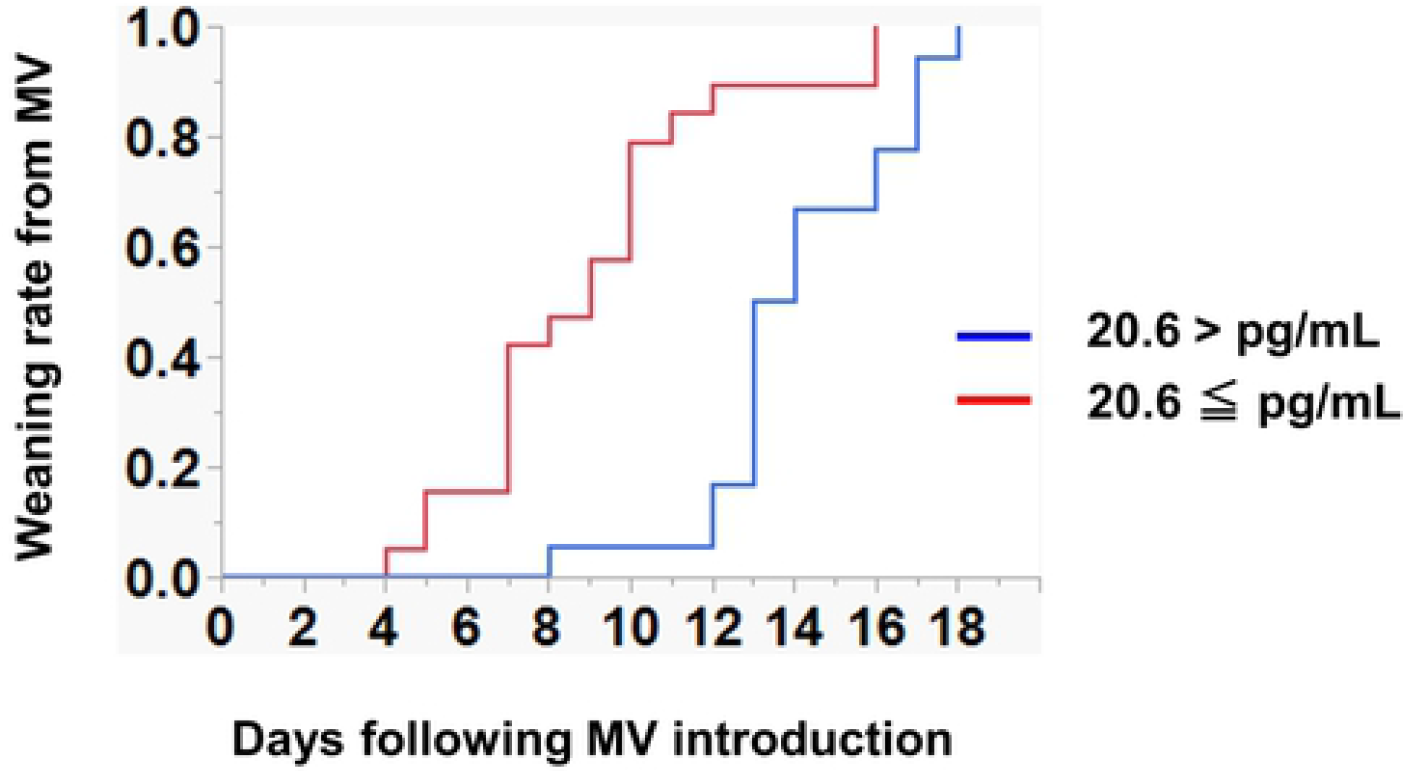
Kaplan-Meier analysis for weaning from invasive mechanical ventilation (MV) Kaplan-Meier curves showing the cumulative weaning rate from invasive MV in SARS-CoV-2-mediated ARDS patients who were treated with dexamethasone according to IL-6 levels above or below the optimal cutoffs (20.6 pg/mL) on day 7 after intubation.

## Discussion

So far, IL-6 on admission or before intubation generated an interest as a predictor of developing ARDS. Elevation of IL-6 levels in plasma has been related with the risk of requiring ICU management, MV, or ECMO in patients with COVID-19.^25-30^ Recently, Awasthi et al associated plasma level of IL-6 with the duration of ICU stay in COVID-19 patients.^31^ Moreover, an observational study reported that IL-6 level predicts requirement of invasive MV and indication for the blockade of IL-6 receptor with tocilizumab in COVID-19.^32^ However, the clinical utility of measuring IL-6 level after intubation and in patients with established ARDS has not been elucidated. The present study extends the emerging clinical utility of IL-6 to predict early withdrawal from MV in intubated patients with ARDS caused by SARS-CoV-2. A subset of survivors from established ARDS has been proven to develop irreversible fibrotic changes in disease lung, including fibroblast accumulation, deposition of collagen and other lung extracellular matrix components, which leads to high mortality.^33^ Thille et al demonstrated that the prevalence of fibrotic change in pulmonary origin is closely related with ARDS duration.^34^ Moreover, although the MV has facilitated the management of ARDS, ventilator-induced lung injury has been shown to be a predominant causative factor for lung fibrosis in ARDS patients.^35^ Thus, early withdrawal from MV is an important therapeutic strategy for established ARDS. Recently, efficacy of steroid administration on MV duration in established ARDS has been reported.^10^ Patients with severe ARDS have been shown to frequently require MV, with a mean duration of approximately 12 days, when they were treated with steroids.^36^

Consistently, a recent randomized controlled trial of COVID-19 in patients with ARDS showed that treatment with dexamethasone significantly shortens the duration of MV use (a mean duration of 12.5 vs. 13.9 days).^37^ Our data also supported this beneficial effect (a mean MV duration of 11.5 vs. 16.1 days). However, no indicators have been identified to predict responsiveness to dexamethasone. Importantly, we are the first to show that IL-6 levels serve as a predictive indictor of earlier MV withdrawal after dexamethasone administration in invasively ventilated patients with SARS-CoV-2-induced ARDS, and that patients with IL-6 levels below or above 20.6 pg/mL on day 7 after intubation are likely to withdraw from MV within 11 days or to require MV more persistently, respectively. Accordingly, measuring IL-6 may help monitoring therapeutic efficacy and facilitate steroid therapy in invasively ventilated patients with established ARDS and contribute to the improvement of short- and long-term outcomes by early withdrawal from MV and mitigation or prevention of lung fibrosis. Moreover, these observations also provide a warrant for further investigation of the potential IL-6-guided therapy for established ARDS. Capturing trends in IL-6 levels over time may be useful for understanding the clinical course in COVID-19. For example, elevation of consecutive IL-6 concentrations shows a predictive value for changes in disease severity from moderate to severe or critical. ^38^ Moreover, the re-elevation of IL-6 level after MV introduction or during treatment with steroid may be indicative of ventilator-induced lung injury and secondary bacterial infections, respectively.^39-40^ If so, neuromuscular blocking agents to suppress excessive spontaneous-breathing efforts or antibiotics for secondary pneumonia will be needed. Our data also associated changes in IL-6 levels after intubation with earlier withdrawal from invasive MV in SARS-CoV-2-induced ARDS. However, we could not confirm that the 2-day difference and percentage of change in IL-6 have better accuracy than the absolute level, because in the present study, some patients were started dexamethasone administration, which decreases baseline levels of IL-6, prior to admission to our institution. Analysis in a patient population with standardized steroid initiation times may improve their accuracies. Clinical utility of changes in IL-6 levels should be investigated.

In the present data, absolute levels and percentage of change in CRP were also associated with earlier withdrawal from invasive MV. Due to the small sample size, it was not possible to statistically compare the predicted values of CRP and IL-6. However, synthesis and release of CRP protein, which is widely used as a biomarker for inflammatory status, from liver and immune cells depends upon stimulation by IL-6.^41-42^ This supports the superiority of IL-6 in early predicting disease activity of established ARDS. Indeed, a previous study demonstrated that IL-6 level can earlier predict the requirement of MV in severe COVID-19 compared with CRP level.^29^

The present study has several strengths: First, the present results showed that IL-6 can predict not only the risk of developing ARDS, but also therapeutic response, such as early withdrawal from invasive MV, in established ARDS, which may suggest a potential utility as a determinant of therapeutic strategy, including continuation of treatment and the need for additional treatment. Second, it is easy to measure circulating IL-6 level over time in various clinical settings, as instruments for simple and rapid measurement are already widely available.^43-44^ Third, IL-6 has been widely recognized as a prognostic indicator of inflammatory diseases such as sepsis ^45^ and rheumatoid arthritis,^46^ making it easy for medical staff to interpret. Finally, IL-6 in plasma may be available in evaluating and monitoring the therapeutic effect of new drugs on inflammatory diseases including ARDS.

There are several limitations of this study. The data set of this study was a backward-looking study, so there were some missing values. Second, this study was conducted at a single institution, resulting in a biased patient population. Third, our sample size is too small to evaluate optimal cutoffs of variables and to determine clinical utility of IL-6 in the treatment of ARDS and its superiority over other markers. Fourth, the cause of ARDS was limited to SARS-CoV-2 infection. Fifth, steroid therapy was not standardized in our study population. Thus, the applicability of IL-6 in clinical practice needs to be prospectively studied in large cohorts of strictly steroid-treated patients with ARDS derived from various etiologies.

## Conclusions

We are the first to present data illustrating that IL-6 in plasma is a promising indicator for predicting earlier withdrawal from invasive MV after dexamethasone administration in intubated patients with SARS-CoV-2-mediated ARDS. This suggests that measuring IL-6 may allow monitoring responsiveness to dexamethasone and help determine therapeutic strategy in established ARDS. Our data provide the first evidence suggesting the importance of measuring IL-6 level after intubation and in patients with established ARDS.

## Data Availability

All relevant data are within the manuscript and its Supporting Information files.

## List of abbreviations

ARDS: acute respiratory distress syndrome
COVID-19: A new coronavirus
SARS-CoV-2: severe acute respiratory syndrome coronavirus 2
IL: interleukin
CRP: C-reactive protein
MV: mechanical ventilation
ECMO: extracorporeal membranous oxygenation

## Acknowledgments

We thank our colleagues in the Department of Emergency Medicine, Yokohama City University and, the clinical nurses in the Advanced Care Unit, Yokohama City University Hospital for their kind assistance.

## References

1. B. Lium. Adult respiratory distress syndrome (ARDS). Incidence, clinical findings, pathomorphology and pathogenesis. A review. Nord Vet Med. 1983 Jan;35 (1): 38–47.

2. D.R. Janz, L.B. Ware. Biomarkers of ALI/ARDS: pathogenesis, discovery, and relevance to clinical trials. Semin Respir Crit Care Med. 2013 Aug;34 (4): 537–548.

3. D.R. Janz, L.B. Ware. The role of red blood cells and cell-free hemoglobin in the pathogenesis of ARDS. J Intensive Care. 2015 Jun;3 (20).

4. M. Moss. Mortality is the only relevant outcome in ARDS: yes. Intensive Care Med. 2015 Jan; 41(1): 141–143.

5. G. Bellani, J.G. Laffey, T. Pham, E. Fan, L. Brochard, A. Esteban, et al. Epidemiology, Patterns of Care, and Mortality for Patients With Acute Respiratory Distress Syndrome in Intensive Care Units in 50 Countries. JAMA. 2016 Feb;315 (8): 788–800.

6. G.U. Meduri, A.S. Headley, E. Golden, S.J. Carson, R.A. Umberger, T. Kelso, et al. Effect of prolonged methylprednisolone therapy in unresolving acute respiratory distress syndrome: a randomized controlled trial. JAMA. 1998 Jul; 280 (2):159–165.

7. G.U. Meduri, E. Golden, A.X. Freire, E. Taylor, M. Zaman, S.J. Carson, et al. Methylprednisolone infusion in early severe ARDS: results of a randomized controlled trial. Chest. 2007 Apr;131 (4): 954–963.

8. K.P. Steinberg, L.D. Hudson, R.B. Goodman, C.L. Hough, P.N. Lanken, R. Hyzy, Efficacy and safety of corticosteroids for persistent acute respiratory distress syndrome. N Engl J Med. 2006 Apr;354 (16):1671–1684.

9. S. Tongyoo, C. Permpikul, W. Mongkolpun, V. Vattanavanit, S. Udompanturak, M. Kocak, et al. Hydrocortisone treatment in early sepsis-associated acute respiratory distress syndrome: results of a randomized controlled trial. Crit Care. 2016 Oct;20 (1):329.

10. J. Villar, C. Ferrando, D. Martínez, A. Ambrós, T. Muñoz, J.A. Soler. Dexamethasone treatment for the acute respiratory distress syndrome: a multicentre, randomised controlled trial. Lancet Respir Med. 2020 Mar;8 (3):267–276.

11. J.T. Wu, K. Leung, G.M. Leung. Nowcasting and forecasting the potential domestic and international spread of the 2019-nCoV outbreak originating in Wuhan, China: A modelling study. Lancet. 2020 Feb;395 (10225):689–697.

12. X. Yang, Y. Yu, J. Xu, H. Shu, J. Xia, H. Liu, et al. Clinical course and outcomes of critically ill patients with SARS-CoV-2 pneumonia in Wuhan, China: a single-centered, retrospective, observational study. Lancet Respir Med. 2020 May;8 (5):475–481.

13. J.H. Beigel, K.M. Tomashek, L.E. Dodd, A.K. Mehta, B.S. Zingman, A.C. Kalil, et al. Remdesivir for the Treatment of Covid-19 - Final Report. N Engl J Med. 2020 Nov;383 (19):1813–1826.

14. P. Horby, W.S. Lim, J.R. Emberson, M. Mafham, J.L. Bell, L. Linsell, et al. Dexamethasone in Hospitalized Patients with Covid-19—Preliminary Report. N Engl J Med. 2021 Feb;384 (8).693–704.

15. S. Tasaka, A. Ishizaka, T. Urano, K. Sayama, F. Sakamaki, H. Nakamura, et al. BCG priming enhances endotoxin-induced acute lung injury independent of neutrophils. Am J Respir Crit Care Med. 1995 Sep;152 (3):1041–1049.

16. C. Song, H. Li, Y. Li, M. Dai, L. Zhang, S. Liu, et al. NETs promote ALI/ARDS inflammation by regulating alveolar macrophage polarization. Exp Cell Res. 2019 Sep;382 (2):111486.

17. C. Wang, J. Xie, L. Zhao, X. Fei, H. Zhang, Y. Tan, et al. Alveolar macrophage dysfunction and cytokine storm in the pathogenesis of two severe COVID-19 patients. EBioMedicine. 2020 Jul;57:102833.

18. T.R. Martin. Lung cytokines and ARDS: Roger S. Mitchell Lecture. Chest. 1999 Jul;116 (1 Suppl):2S–8S.

19. H. Schütte, J. Lohmeyer, S. Rosseau, S. Ziegler, C. Siebert, H. Kielisch, et al. Bronchoalveolar and systemic cytokine profiles in patients with ARDS, severe pneumonia and cardiogenic pulmonary oedema. Eur Respir J. 1996 Sep;9 (9):1858–1867.

20. G.U. Meduri, S. Headley, G. Kohler, F. Stentz, E. Tolley, R. Umberger, et al. Persistent elevation of inflammatory cytokines predicts a poor outcome in ARDS. Plasma IL-1 beta and IL-6 levels are consistent and efficient predictors of outcome over time. Chest. 1995 Apr;107 (4):062–1073.

21. M. Liao, Y. Liu, J. Yuan, Y. Wen, G. X, J. Zhao, et al. Single-cell landscape of bronchoalveolar immune cells in patients with COVID-19. Nat Med. 2020 Jum;26 (6).;842–844.

22. A.J. Wilk, A. Rustagi, N.Q. Zhao, J. Roque, G.J. Martínez-Colón, J.L. McKechnie, et al. A single-cell atlas of the peripheral immune response in patients with severe COVID-19. Nat Med. 2020 Jul;6 (7) (2020):1070–1076.

23. V.M. Ranieri, G.D. Rubenfeld, B.T. Thompson, N.D. Ferguson, E. Caldwell, E. Fan, et al. Acute respiratory distress syndrome: The Berlin definition. JAMA. 2012 Jul;307 (23):2526–2533.

24. S. Hashimoto, M. Sanui, M. Egi, S. Ohshimo, J. Shiotsuka, R. Seo, R. Tanaka, et al. The clinical practice guideline for the management of ARDS in Japan. J Intensive Care. 2017 July;5 (50):50.

25. D.M. Del Valle, S. Kim-Schulze, H.H. Huang, N.D. Beckmann, S. Nirenberg, B. Wang, et al. An inflammatory cytokine signature predicts COVID-19 severity and survival. Nat Med. 2020 Oct;26(10) ;1636–1643.

26. A. Vultaggio, E. Vivarelli, G. Virgili, E. Lucenteforte, A. Bartoloni, C. Nozzoli, et al. Prompt Predicting of Early Clinical Deterioration of Moderate-to-Severe COVID-19 Patients: Usefulness of a Combined Score Using IL-6 in a Preliminary Study. J Allergy Clin Immunol Pract. 2020 Sep;8(8):2575–2581.e2.

27. R. Laguna-Goy, A. Utrero-Rico\, P. Talayero, M. Lasa-Lazaro, A. Ramirez-Fernandez, L. Naranjo, et al. IL-6–based mortality risk model for hospitalized patients with COVID-19. J Allergy Clin Immunol. 2020 Oct;146 (4):799–807.e.9.

28. R. Saji, M. Nishii, K. Sakai, K. Miyakawa, Y. Yamaoka, T. Ban, et al. Combining IL-6 and SARS-CoV-2 RNAaemia-based risk stratification for fatal outcomes of COVID-19. PLoS One. 2021 Aug;16 (8);.e0256022.

29. T. Herold, V. Jurinovic, C. Arnreich, B.J. Lipworth, J.C. Hellmuth, M.V. Bergwelt-Baildon, et al. Elevated levels of IL-6 and CRP predict the need for mechanical ventilation in COVID-19. J Allergy Clin Immunol.2020 Jul;146 (1):128–136.e.4.

30. S. Keddie, O. Ziff, M.K.L. Chou, R.L. Taylor, A. Heslegrave, E. Garr, et al. Laboratory biomarkers associated with COVID-19 severity and management. Clin Immunol. 2020 Dec;221:108614.

31. S. Awasthi, T. Wagner, A.J. Venkatakrishnan, A. Puranik, M. Hurchik, V. Agarwal, et al. Plasma IL-6 levels following corticosteroid therapy as an indicator of ICU length of stay in critically ill COVID-19 patients. Cell Death Discov. 2021 Mar;7 (1):55.

32. J.M. Galván-Román, S.C. Rodríguez-García, E. Roy-Vallejo, A. Marcos-Jiménez, S. Sánchez-Alonso, C. Fernández-Díaz, et al. IL-6 serum levels predict severity and response to tocilizumab in COVID-19: An observational study. J Allergy Clin Immunol. 2021 Jan;147 (1):72-80.e8.

33. E.L. Burnham, W.J. Janssen, D.W. Riches, M. Moss, G.P. Downey. The Fibroproliferative Response in Acute Respiratory Distress Syndrome: Mechanisms and Clinical Significance. Eur Respir J. 2014 Jan;43 (1):276–285.

34. A.W. Thille, A. Esteban, P. Fernández-Segoviano, J.M. Rodriguez, J.A. Aramburu, P. Vargas-Errázuriz, et al. Chronology of Histological Lesions in Acute Respiratory Distress Syndrome With Diffuse Alveolar Damage: A Prospective Cohort Study of Clinical Autopsies. Lancet Respir Med. 2013 Jul;1(5):395–401.

35. N.E. Cabrera-Benitez, J.G. Laffey, M. Parotto, P.M. Spieth, J. Villar, H. Zhang, et al. Mechanical Ventilation-Associated Lung Fibrosis in Acute Respiratory Distress Syndrome: A Significant Contributor to Poor Outcome. Anesthesiology. 2014 Jul;121 (1):189–198.

36. G.U. Meduri, P.R. Rocco, D. Annane, S.E. Sinclair. Prolonged glucocorticoid treatment and secondary prevention in acute respiratory distress syndrome. Expert Rev Respir Med. 2010 Apr;4 (2) :201–210.

37. B.M. Tomazini, I.S. Maia, A.B. Cavalcanti, O. Berwanger, R.G. Rosa, V.C. Veiga, et al. Effect of Dexamethasone on Days Alive and Ventilator-Free in Patients With Moderate or Severe Acute Respiratory Distress Syndrome and COVID-19: The CoDEX Randomized Clinical Trial. JAMA. 2020 Oct;324 (13):1307–1316.

38. X. Chen, J. Zhou, C. Chen, B. Hou, A. Ali, F. Li, et al. Consecutive Monitoring of Interleukin-6 Is Needed for COVID-19 Patients. Virol Sin. 2021;36 (5):1093–1096.

39. A.S. Slutsky. Neuromuscular blocking agents in ARDS. N Engl J Med. 2010 Sep;363 (12):1176–1180.

40. R. Obata, T. Maeda, D. Rizk, T. Kuno. Increased Secondary Infection in COVID-19 Patients Treated with Steroids in New York City. Jpn J Infect Dis. 2021 Jul;74 (4):307–315.

41. J.V. Castell, M.J. Gómez-Lechón, M. David, T. Andus, T. Geiger, R. Trullenque, et al. Interleukin-6 is the major regulator of acute phase protein synthesis in adult human hepatocytes. FEBS Lett. 1989 Jan;242 (2):237–239.

42. P.B. Sehgal. Interleukin-6: a regulator of plasma protein gene expression in hepatic and non-hepatic tissues. Mol Biol Med. 1990 Apr;7 (2):117–130.

43. S.K. Fischer, K. Williams, L. Wang, E. Capio, M. Briman. Development of an IL-6 point-of-care assay: utility for real-time monitoring and management of cytokine release syndrome and sepsis. Bioanalysis. 2019 Oct;11 (19):1777–1785.

44. D.W. Jekarl, S.Y. Lee, J. Lee, Y.J. Park, Y. Kim, J.H. Park. Procalcitonin as a diagnostic marker and IL-6 as a prognostic marker for sepsis. Diagn Microbiol Infect Dis. 2013 Apr;75 (4):342–347.

45. P. Damas, D. Ledoux, M. Nys, Y. Vrindts, D. De Groote, P. Franchimont, M. Lamy. Cytokine serum level during severe sepsis in human IL-6 as a marker of severity. Ann Surg. 1992 Apr 215 (4) :356–362.

46. J.E. Gottenberg, J.M. Dayer, C. Lukas, B. Ducot, G. Chiocchia, A. Cantagrel, et al. Serum IL-6 and IL-21 are associated with markers of B cell activation and structural progression in early rheumatoid arthritis: results from the ESPOIR cohort. Ann Rheum Dis. 2012 Jul;71 (7):1243–1248.

